# Increased interhemispheric asymmetry of cerebral blood flow in comatose adult patients on extracorporeal membrane oxygenation

**DOI:** 10.1101/2021.10.25.21265432

**Authors:** Thomas W. Johnson, Irfaan Dar, Kelly Donohue, Yama Y. Xu, Esmeralda Santiago, Olga Selioutski, Mark A. Marinescu, Ross K. Maddox, Tong Tong Wu, Giovanni Schifitto, Igor Gosev, Regine Choe, Imad Khan

**Affiliations:** Department of Neurology, University of Rochester Medical Center, Rochester, NY, USA; Department of Biomedical Engineering, University of Rochester, Rochester, NY, USA; Department of Neurology, Northwestern University Feinberg School of Medicine, Chicago, IL, USA; Department of Philosophy, University of Rochester, Rochester, NY, USA; Department of Medicine, University of Rochester Medical Center, Rochester, NY, USA; Department of Biostatistics and Computational Biology, University of Rochester, Rochester, NY, USA; Division of Cardiac Surgery, Department of Surgery, University of Rochester Medical Center, Rochester, NY, USA; Department of Electrical and Computer Engineering, University of Rochester, Rochester, NY, USA

**Keywords:** cerebrovascular autoregulation, coma, diffuse correlation spectroscopy, extracorporeal membrane oxygenation, cerebral blood flow

## Abstract

Extracorporeal membrane oxygenation (ECMO) is a mechanical circulatory support method that is associated with a high burden of neurologic injury, but neurologic examination and imaging of this population presents prohibitive medical and logistical challenges. Diffuse correlation spectroscopy (DCS) can measure relative cerebral blood flow (rBF) non-invasively at the bedside. In this study we observed interhemispheric differences in rBF in response to mean arterial pressure (MAP) changes in adult ECMO recipients. Thirteen patients were recruited (ages 21-78, 7 with cardiac arrest, 4 with acute heart failure, 2 with acute respiratory distress syndrome). They were dichotomized via Glasgow Coma Scale Motor score (GCS-M) into comatose (GCS-M ≤ 4; n=4) and non-comatose (GCS-M > 4; n=9) groups. Comatose patients had more interhemispheric rBF asymmetry (ASYM_rBF_) vs. non-comatose patients over a range of MAP values (29% [IQR 23-34%] vs. 11% [IQR 8-13%], p=0.009). ASYM_rBF_ in comatose patients resolved near a MAP range of 70-80 mm Hg, while rBF remained symmetric through a wider MAP range in non-comatose patients. Associations between post-oxygenator pCO_2_ or pH vs. ASYM_rBF_ individually did not meet significance, though the linear model slopes were different between comatose and non-comatose subgroups. Here we have demonstrated asymmetric cerebral autoregulation in comatose ECMO patients.

## Introduction

Extracorporeal membrane oxygenation (ECMO) is a method of external mechanical circulatory support (MCS) that can both oxygenate and circulate blood, thus supplementing the function of both the lungs and the heart during critical illness. Its use has increased in the preceding decade as technology and techniques have improved. However, approximately 13% of ECMO recipients are found to have neurologic injuries either from underlying pathology or the ECMO therapy itself^1-3^. Performing accurate neurologic examinations in this population is challenging due to high sedation and neuromuscular blockade requirements to prevent cannula dislodgement, optimize hemodynamics, or to force ventilator compliance^4, 5^. Neuroimaging requires resource-intensive, multi-disciplinary transportation to imaging suites^6^, while invasive neuromonitoring can further increase the risk of cerebral hemorrhage in this coagulopathic population^7^. Thus, the brain’s perfusion and oxygenation requirements are not routinely monitored in clinical practice, furthering the potential for injury.

Diffuse correlation spectroscopy (DCS) provides non-invasive, continuous bedside neuromonitoring using near-infrared light, modeled with the correlation diffusion equation^8^, to measure blood flow index (BFI), which is directly proportional to cerebral blood flow (CBF)^9^. Its use to monitor CBF has been described in a number of brain-injured cohorts^10-12^and has been verified against other CBF measurement modalities^13-15^. DCS is closely related to near-infrared spectroscopy (NIRS), a commercially available technology that measures cerebral oxygenation, but DCS offers the advantage of measuring CBF directly rather than using oximetry as a surrogate.

Veno-arterial (VA) ECMO is often configured to supply oxygenated blood in retrograde flow up the aorta via a cannula placed in the femoral artery. This is contrasted with veno-venous (VV) ECMO, in which oxygenated blood is returned to the body via the venous system and perfuses the arterial system in a normal physiologic manner. This distinction is important as asymmetric cerebral and somatic oxygenation and perfusion have been described in VA ECMO^16-18^. The presence of disrupted cerebrovascular autoregulation in comatose patients with hypoxic brain injury furthers the possibility of asymmetric perfusion arising from the brain’s interaction with the ECMO circuit and the unique changes in circulatory physiology ECMO produces^19^. This study uses DCS to observe hemispheric CBF in adult ECMO patients with the hypothesis that comatose patients will exhibit increased interhemispheric asymmetry than in non-comatose patients.

## Material and Methods

### Patient Recruitment

This study was approved by the Institutional Review Board (IRB) at the University of Rochester. Adult patients aged 18 years and older who were admitted to the cardiac intensive care unit (CICU) for ECMO therapy at the University of Rochester Medical Center were eligible for this study. Patients undergoing VA or VV ECMO for any etiology were included. Exclusion criteria consisted of pre-existing neurologic conditions and facial injuries impeding DCS measurement. Informed consent was obtained from patients’ legally authorized representatives 24 hours after ECMO initiation.

### Experimental Design

A detailed description of the experimental design and instrumentation used for this study has been previously published^20^. In summary, enrolled subjects underwent a protocol of daily CBF monitoring using an in-house diffuse correlation spectroscopy (DCS) system. The DCS system employs a 785 nm laser with long coherence length to non-invasively quantify blood flow index (BFI), which is proportional to CBF^13-15, 21^. Two slim-profiled optical probes using a 45-degree prism were attached to the left and right forehead of the patient using double-sided tape (3M, St. Paul, MN) and Tegaderm (3M, St Paul MN). Light emitted from these two probes propagated into the tissue, then was collected by single mode optical fibers at 2.5 cm separation from the source that were connected to single-photon counting detectors. Autocorrelation curves were calculated using a 2 second integration time, which translated to BFI values at 0.25 Hz for each hemisphere. BFI values were calculated from the autocorrelation curves using constant absorption and scattering coefficients previously reported in the literature for brain measurements^20, 22-24^. Data was discarded if artifacts were present or had poor signal to noise ratio (SNR), which was defined as less than 4 kilo-counts per second (kcps) measured by the detectors.

Continuous physiological data including systolic, diastolic, and mean arterial pressure (MAP) were recorded continuously at the bedside using Medicollector software (Medicollector Inc, Boston, MA) at 1 Hz. ECMO pump speed and flow were also recorded at 0.2 Hz from the ECMO machine. BFI and physiologic monitoring were performed for up to 3 hours during the initial resuscitation phase, during which ECMO fully supported the circulation at constant pump speeds. Monitoring was performed for up to 8 hours during the ECMO wean phase as pump speeds were gradually tapered by clinical staff to test native cardiovascular function. This prolonged duration was performed to capture as much hemodynamic change as possible.

### Cerebral Autoregulation Asymmetry Analysis

Cerebral autoregulation curves correlating cerebral perfusion with mean arterial pressure (MAP) were created from left and right hemispheric DCS data for all patients. All analysis was performed using MATLAB 2020 (Mathworks, Natick, MA). BFI and MAP data were first resampled to 0.25 Hz to ensure a uniform time series between the data sets. The continuous measurement of relative blood flow (rBF, in %) was determined for each day of DCS monitoring to evaluate the relative change in BFI for all subjects. This was calculated by normalizing the BFI data of each hemisphere by the median BFI value of that hemisphere^22^. Figure 1a displays the rBF for the right hemisphere of one subject while Figure 1b shows the concurrent MAP. Using the concurrent rBF and MAP time series, rBF values were plotted against MAP values between 50 and 120 mmHg at 1 mmHg increments shown in the background scatterplot of Figure 1c. From this scatterplot, the average rBF value was calculated at each MAP value, shown as the red circles in Figure 1c. Average rBF values were discarded if there were less than 5 data points at the specific MAP value. Figure 1d shows the average curve for both left and right hemisphere.

**Figure 1:**
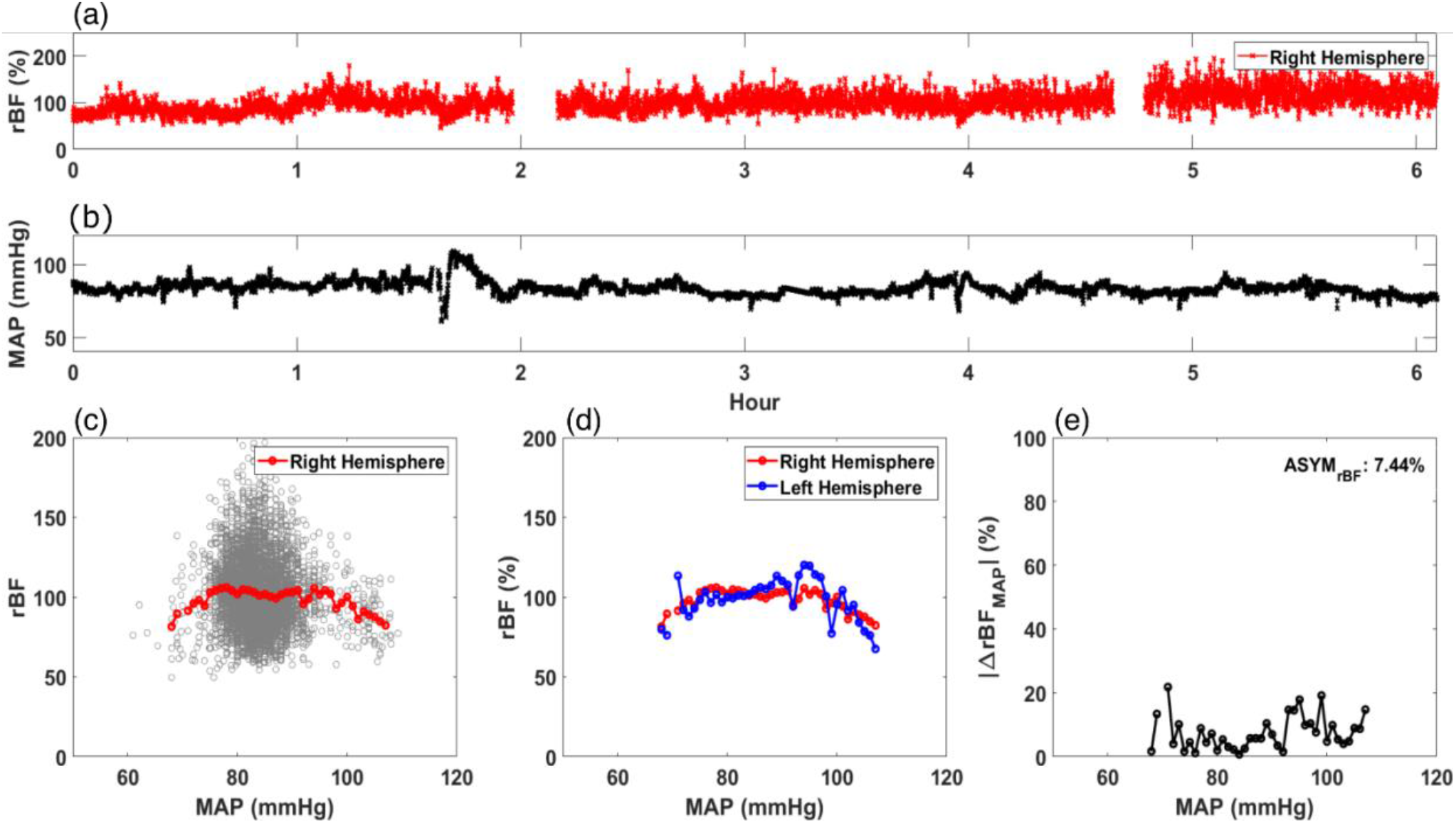
Calculation of average relative blood flow (rBF, in %) versus mean arterial pressure (MAP). An example time trace for (a) rBF and (b) MAP for a single day of monitoring are shown. Using these time traces, (c) rBF is plotted against MAP and an average rBF value at each MAP is calculated (red circle). (d) An averaged rBF vs MAP curve is generated for both the right (red circle) and left hemisphere (blue circle). Using the data in (d), |ΔrBF_MAP_| (in %/mmHg) is plotted against MAP in (e). ASYM_rBF_, averaged |ΔrBF_MAP_| of this monitoring day for a patient is shown on the upper right corner.

Using these data, asymmetry between the hemispheric data was calculated for each day of monitoring for all subjects. First, the magnitude of difference was calculated as |ΔrBF_MAP_(MAP_i_)| defined in Equation 1 and reported as %. MAP_i_ are the MAP values in increments of 1 mmHg derived from the MAP range that day, to a maximum MAP value at MAP_N_, where N is the total number of MAP increments of the monitoring day. We averaged |ΔrBF_MAP_(MAP_i_)| to calculate ASYM_rBF_ for the monitoring day, shown in Equation 2 and Figure 1e, reported as %.

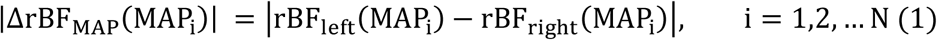

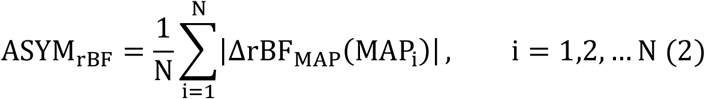

### Clinical Characteristics

Data obtained from the electronic medical record data for each subject were recorded in a secure REDCap database. We recorded demographics, ECMO characteristics (duration, location of cannula, reason for cannulation), and daily clinical characteristics. Clinical characteristics included Glasgow Coma Scale (GCS), Sequential Organ Failure Assessment (SOFA), arterial blood gas (ABG) parameters including pH, partial pressure of arterial oxygen (pO_2_), partial pressure of arterial CO_2_ (pCO_2_), lactate, cardiac left ventricular ejection fraction (LVEF), and sedation/analgesia infusion rates.

### Cerebral Autoregulation Asymmetry vs Comatose Status

Each subject’s GCS motor (GCS-M) subscores were recorded daily during monitoring. Subjects who were able to attain GCS-M > 4 at any time during ECMO were characterized as ‘non-comatose’, while those who remained GCS-M ≤ 4 were categorized as ‘comatose’. To evaluate the difference in ASYM_rBF_ between comatose and non-comatose patients, the maximum value of ASYM_rBF_ from the monitoring period was determined.

### Statistical Analysis

One-tailed and two-tailed two-sample t-tests were used to compare maximum ASYM_rBF_ in comatose versus non-comatose patients. Normalcy was checked, and variances between the two groups were tested using a F-test. Alternative hypotheses were defined for both the one-tailed and two-tailed test. With the F-test result and appropriate alternative hypothesis, p-values were calculated for both the one-tailed and two-tailed t-test. The p-value was compared to a significance level α of 0.05, and was considered statistically significant if less than α. Scatter plots with Pearson correlation coefficients were calculated to assess whether pCO_2_, pO_2_, pH, and LVEF associated with either maximal or daily ASYM_rBF_ for each patient. Statistical analysis was performed using MATLAB software.

## Results

Thirteen subjects undergoing ECMO treatment for different indications were enrolled in this study (five females, mean age 46 years old [range 21-78]). Patient demographics and pertinent clinical parameters are shown in Table 1. Seven subjects required ECMO for cardiac arrest, four for cardiogenic shock and two for acute respiratory distress syndrome (ARDS). VA ECMO was performed on 11 patients and VV ECMO on the two patients with ARDS. The mean length of stay was 27 days. The mean duration of ECMO treatment was 266 hours, or 11 days. Mean pre-ECMO SOFA score was 12. Nine subjects (69%) were classified as “awake” and four (31%) as “comatose” based on GCS-M. Nine subjects survived to ECMO decannulation and seven to discharge. A total of 63 monitoring sessions were conducted, with an average of five sessions per subject. Data from seven sessions were omitted from analysis due to low SNR (subject 3, days one through six and subject 11, day three). Subject 3 had jaundice during the initial days on ECMO which affected DCS signal strength and were discarded. Subject 11 had low signal from the right hemisphere on day three which was discarded.

**Table 1:** Patient demographics and pertinent clinical data. LOS: Length of Stay. SOFA: Sequential Organ Failure Assessment. ARDS: Acute Respiratory Distress Syndrome. GCS-M: Glasgow Coma Scale Motor Score. N/A: Not Applicable

| Subject | Age Range | Sex | ECMO Duration (hours) | LOS (days) | ECMO Indication | ECMO Type | Pre-ECMO SOFA | Best GCS-M | Survival to Discharge | Cause of Death |
| --- | --- | --- | --- | --- | --- | --- | --- | --- | --- | --- |
| 1 | 71-75 | F | 93.2 | 40 | Cardiac arrest | VA | 14 | 6 | Y | N/A |
| 2 | 76-80 | F | 433.6 | 10 | Cardiogenic shock | VA | 10 | 6 | N | Sepsis |
| 3 | 36-40 | M | 321.0 | 19 | Cardiogenic shock | VA | 15 | 6 | N | Hemorrhagic shock |
| 4 | 26-30 | F | 159.2 | 26 | Cardiogenic shock | VA | 12 | 6 | Y | N/A |
| 5 | 51-55 | M | 240.7 | 66 | Cardiac arrest | VA | 9 | 6 | Y | N/A |
| 6 | 71-75 | M | 90.3 | 33 | Cardiac arrest | VA | 12 | 6 | Y | N/A |
| 7 | 26-30 | M | 211.8 | 42 | ARDS | VV | 16 | 6 | Y | N/A |
| 8 | 61-65 | M | 131.6 | 21 | Cardiogenic shock | VA | 13 | 4 | N | Cardiovascular |
| 9 | 61-65 | M | 139.1 | 23 | Cardiac arrest | VA | 11 | 1 | N | Sepsis |
| 10 | 21-25 | M | 817.1 | 84 | ARDS | VV | 12 | 6 | Y | N/A |
| 11 | 21-25 | F | 304.1 | 54 | Cardiac arrest | VA | 13 | 4 | Y | N/A |
| 12 | 41-45 | F | 345.7 | 17 | Cardiac arrest | VA | 13 | 6 | N | Cardiovascular |
| 13 | 56-60 | M | 171.7 | 7 | Cardiac arrest | VA | 12 | 1 | N | Cardiovascular |

Daily GCS-M scores recorded during ECMO treatment for each subject are shown in Figure S1. Subjects 1, 2, and 4 were awake and remained so, while subjects 8, 9, 11, and 13 remained comatose throughout monitoring. GCS-M scores for subjects 3, 5, 7, 10, and 12 fluctuated but were able to improve to follow commands at some point. Notably, subjects 7 and 10 awoke to follow commands after paralytics and sedation were weaned, while the exam for subject 5 improved spontaneously on day four.

Table 2 details the clinical characteristics for all subjects. These include cumulative sedation the subjects received, arterial blood gas results, and ECMO circuit pCO_2_ measured both pre and post-oxygenator on the day of maximum observed ASYM_rBF_. Table S1 shows the mean values of these clinical characteristics for the comatose and non-comatose groups. None of the values were significantly different between the two groups.

**Table 2:**
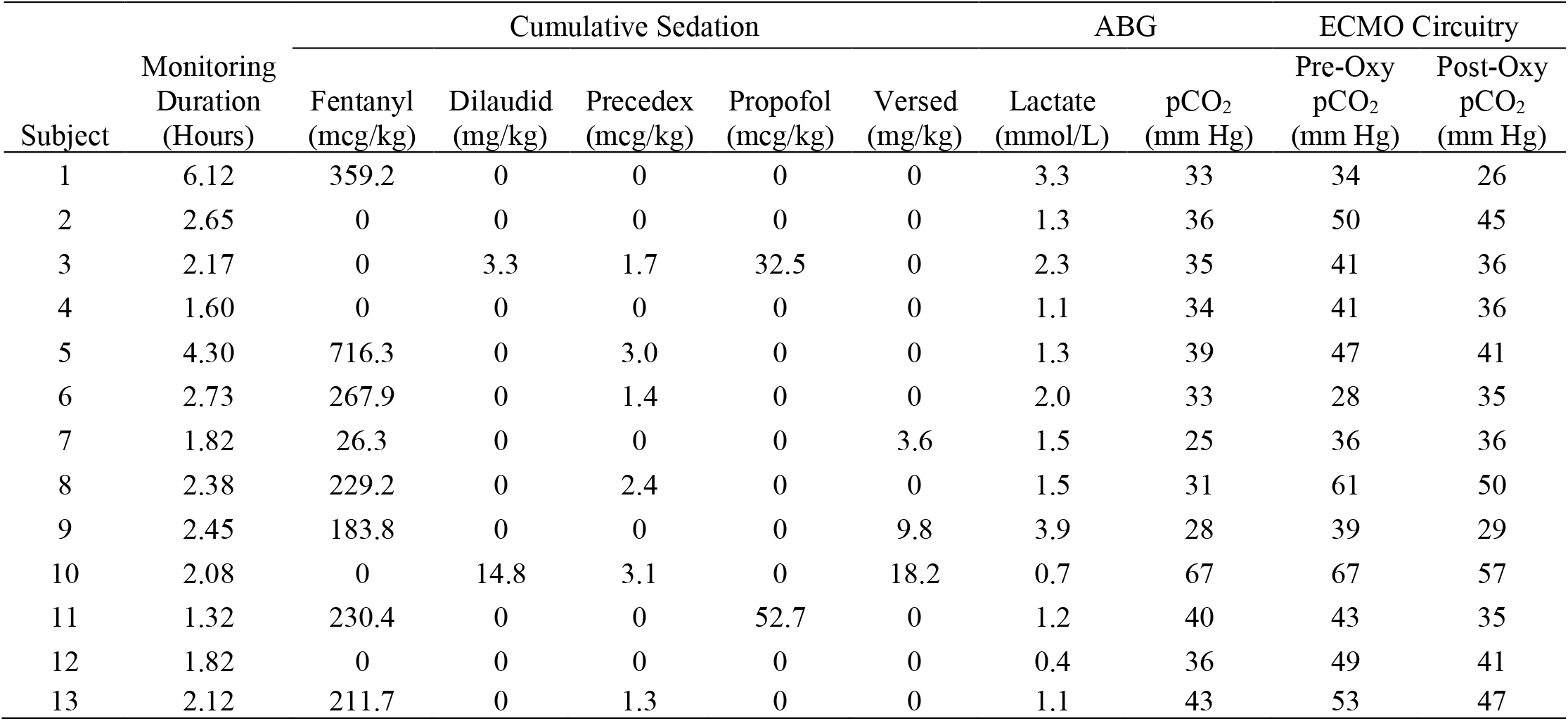
Clinical characteristics of all patients for the day of maximum ASYM_rBF_. Sedation values are the cumulative amount during monitoring period. ABG: Arterial Blood Gas. Pre- and Post-Oxy indicate blood drawn from pre and post oxygenator of the ECMO machine.

As archetypes of the observed asymmetry phenomenon, Figure 2 highlights ASYM_rBF_ vs. GCS-M data for subjects 1 (awake, conversant) and 11 (comatose, severely brain injured, hypoxic-ischemic encephalopathy confirmed by 3 CT scans and 1 MRI scan, due to cardiac arrest). Daily ASYM_rBF_ is shown for all subjects in Figure S2. Figure 3 displays the |ΔrBF_MAP_| for all patients for the specific day that corresponded to their maximum ASYM_rBF_ value highlighted in Figure S2. Comatose subjects experienced significantly higher ASYM_rBF_ values compared to awake subjects (29% [IQR 23-34%] vs. 11% [IQR 8-13%], one-tailed p=0.009, two-tailed p=0.018, Fig. 4).

**Figure 2:**
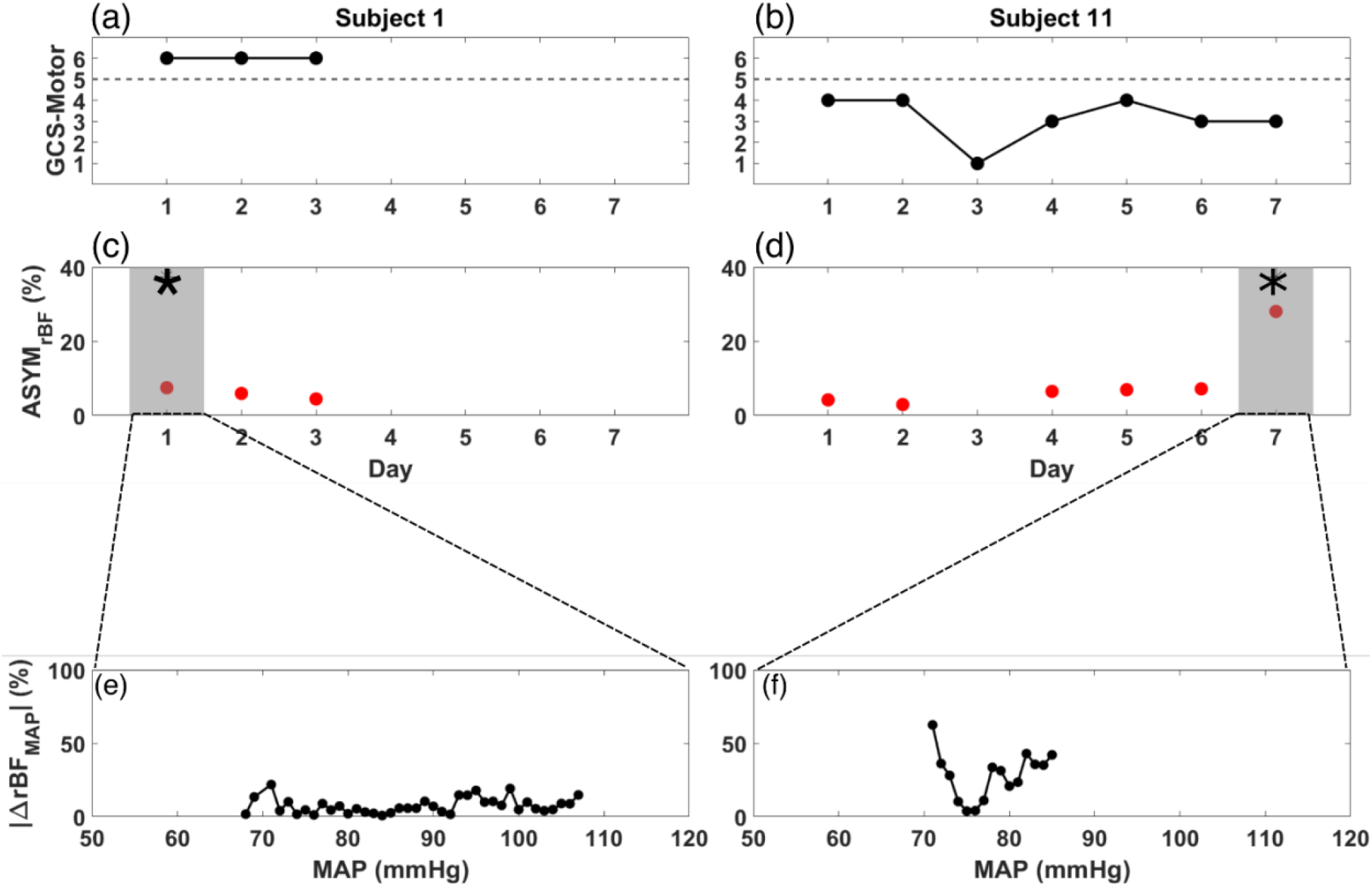
GCS-motor score (black), ASYM_rBF_ (red), and |ΔrBF_MAP_| vs MAP from the day of maximum ASYMrBF for subject 1 (a,c,e) and subject 11 (b,d,f) during their monitoring periods. (a) Subject 1, defined as non-comatose, displayed a constant GCS-motor score of 6 while (b) subject 11, defined as non-comatose, varied between 1 to 4. ASYM_rBF_ data is shown for (c) subject 1 and (d) subject 11. The shaded region marked by the asterisk denotes the maximum ASYM_rBF_ value that was chosen for each subject. |ΔrBF_MAP_| of the gray shaded region shown in (c,d) is shown for (e) subject 1 and (f) subject 11. BFI data for day 3 of subject 11 was discarded due to low SNR thus ASYM_rBF_ was not calculated.

**Figure 3:**
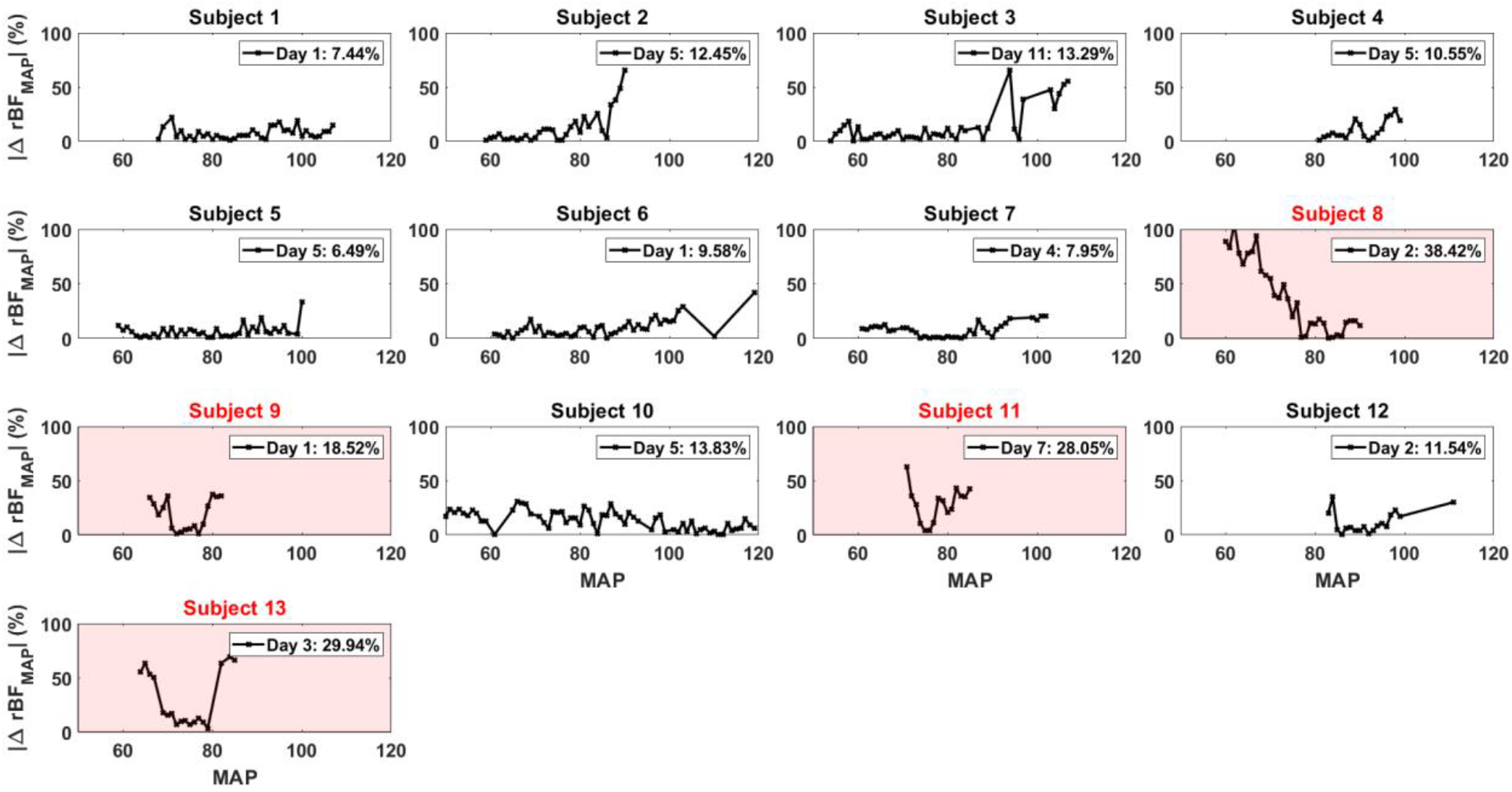
|ΔrBF_MAP_| for all the patients for each specific day that corresponded to their maximum ASYM_rBF_ value. The number displayed in the legend of each plot was the ASYM_rBF_ for that day. Subjects 8, 9, 11, and 13 which are highlighted in red, were subjects defined to be comatose.

**Figure 4:**
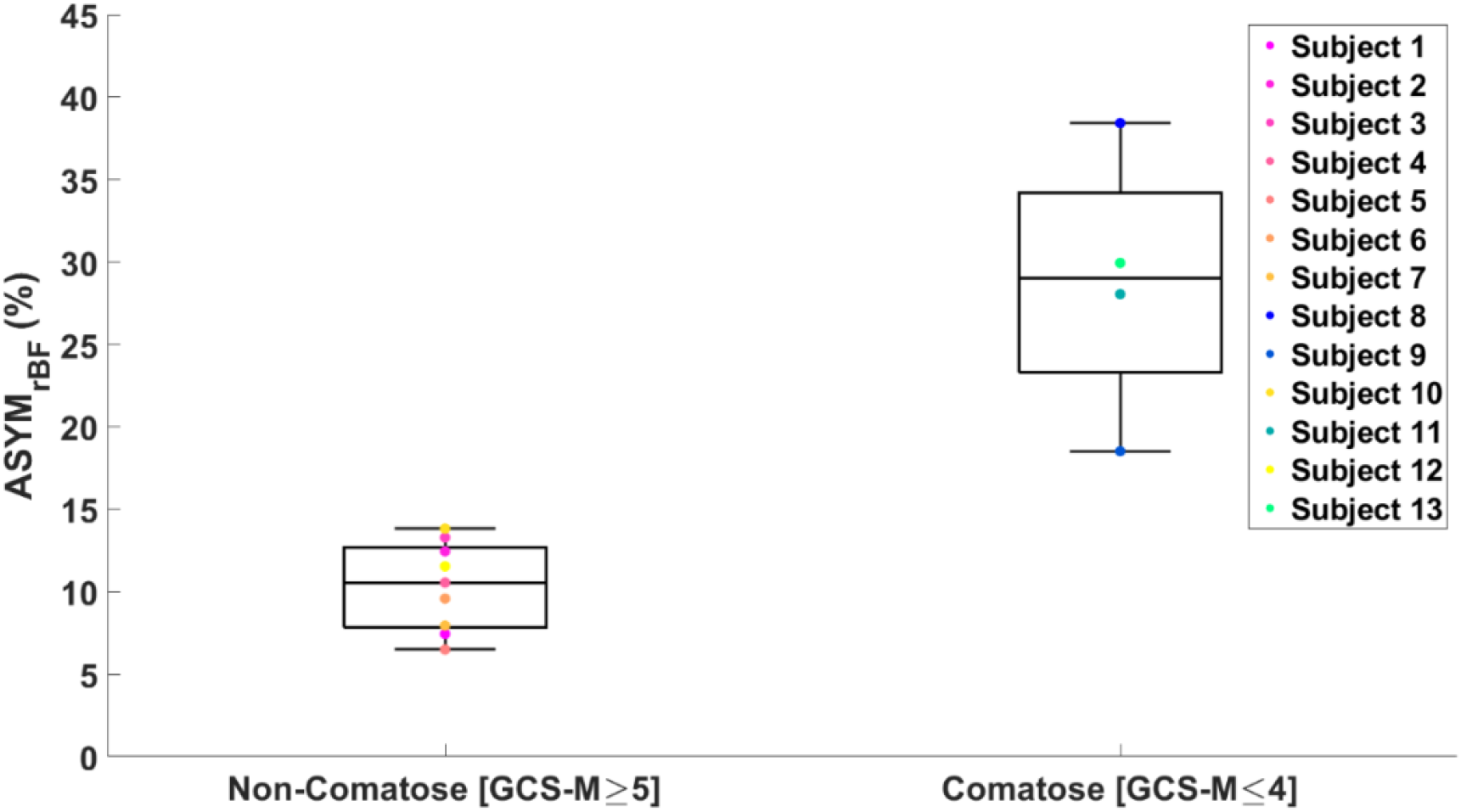
Boxplot of maximum ASYM_rBF_ values for all the subjects grouped by GCS-Motor score. Non-comatose patients had a median value of 11% [IQR 8-13%] and comatose patients 29% [IQR 23-34%] (one-tailed p=0.009, two-tailed p=0.018).

Time-courses of ΔrBF_MAP_ during the total monitoring period on the day of maximum asymmetry is shown for all patients in Figure S3. When focusing on comatose patients shown in red, the direction of asymmetry is of interest. Subject 8 showed rBF skewed towards the right hemisphere at lower MAP values while subject 9, 11, and 13 showed rBF values skewed toward the left hemisphere. However, these patients showed equal rBF of both hemispheres at around a MAP value of 80 mmHg. The non-comatose patients show an equal rBF throughout the entire MAP range compared to the non-comatose patients.

Univariate linear modelling with Pearson correlation analysis was performed with ASYM_rBF_ vs. ABG and post-oxygenator pH, pCO_2_ and pO_2_ values, as well as LVEF. Subset of these analyses are shown in Figure 5, with comatose patients shown in red and non-comatose in blue. There were no significant associations between ASYM_rBF_ and pO_2_ or LVEF in either comatose or non-comatose groups and are not shown. Neither were any associations found to be significant for ABG data: ASYM_rBF_ vs. ABG pCO_2_, comatose slope=0.26 %/mmHg (R=0.22, p=0.78), non-comatose slope=0.12 %/mmHg (R=0.53, p=0.15); ASYM_rBF_ vs. ABG pH, comatose slope=51.04 %/(mmol/L) (R=0.24, p=0.76), non-comatose slope=-2.00 %/(mmol/L) (R=-0.03, p=0.93). In addition, there was no significant association found for post-oxygenator data although excellent R values were noted for some cases: ASYM_rBF_ vs. post-oxygenator pCO_2_, comatose slope=0.75 %/mmHg (R=0.91, p=0.09), non-comatose slope=0.19 %/mmHg (R=0.60, p=0.09); ASYM_rBF_ vs. post-oxygenator pH, comatose slope=-105.27 %/(mmol/L) (R=-0.89, p=0.11), non-comatose slope=-11.87 %/(mmol/L) (R=-0.26, p=0.50). However, comparison of linear model slopes between the comatose and non-comatose subgroups did reveal a significant difference both for post-oxygenator pCO_2_(p=0.03) and post-oxygenator pH (p=0.04).

**Figure 5:**
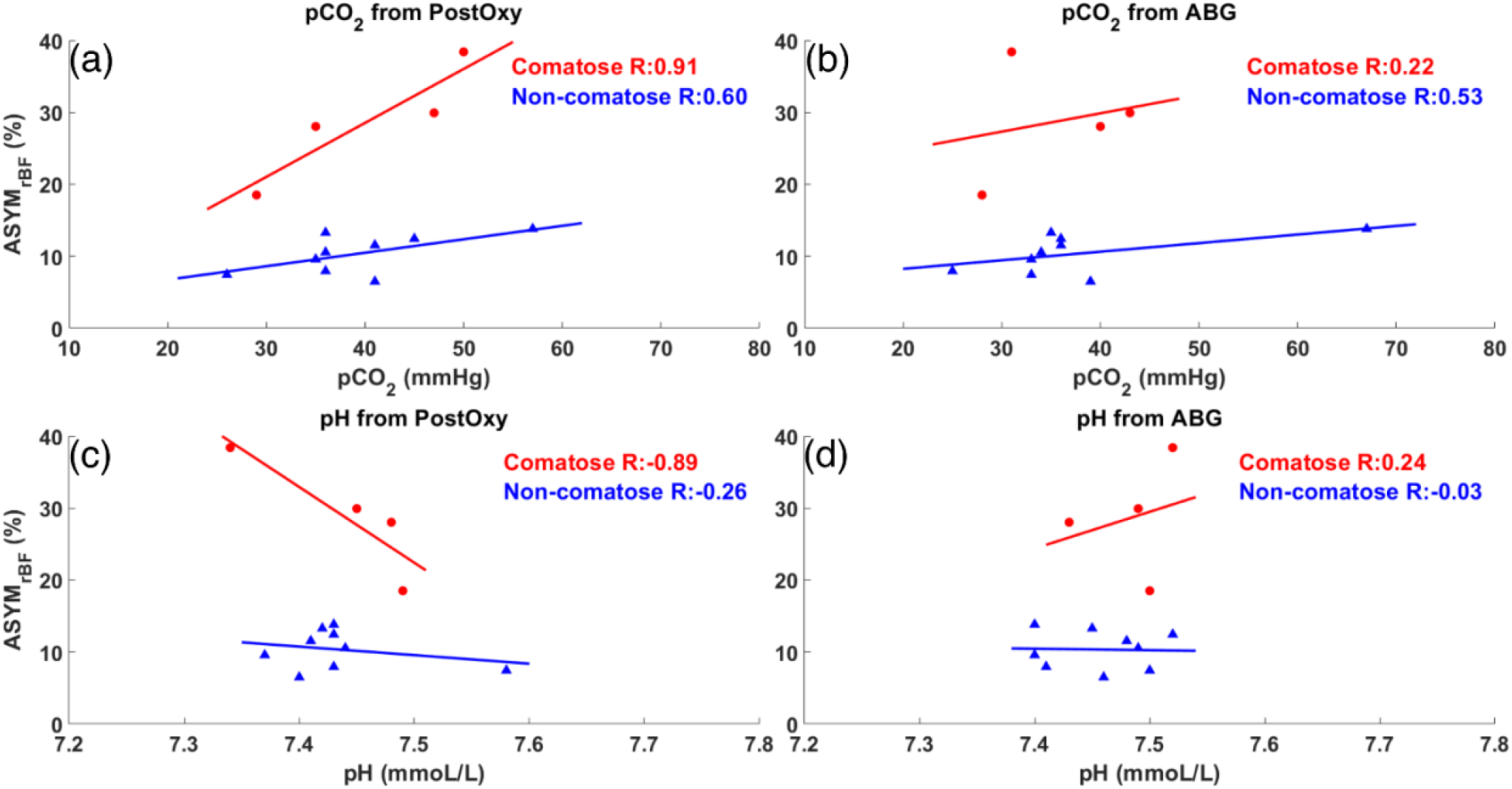
Maximum ASYM_rBF_ vs pCO_2_ and pH values from post-oxygenator and arterial blood gas (ABG). Red circles indicate comatose subjects and blue triangles indicate non-comatose subjects. pCO_2_ and pH values taken from the post-oxygenator are shown in (a,c), while values taken from the ABG are shown in (b,d). Comatose patients showed higher R-value between maximum ASYM_rBF_ and clinical markers (a,c) taken from the post-oxygenator compared to non-comatose patients.

For the non-comatose subjects, all subjects survived to decannulation except subjects 2 and 3. Subject 12 survived decannulation but did not survive until discharge. For the comatose subjects, subjects 8 and 11 survived decannulation, however only subject 11 survived until discharge. Subject 11 was discharged to an acute rehabilitation facility.

## Discussion

In this preliminary study of 13 patients undergoing ECMO, those who remained comatose throughout their duration of treatment were found to have relatively high hemispheric asymmetry in rBF over a range of blood pressures. In these four subjects, the degree of asymmetry appears to change with MAP, but rBF was elevated on the left hemisphere in 75% of them. Though ASYM_rBF_ was found to be elevated in comatose subjects, this metric did not associate with survival to discharge. To our knowledge, this is the first study to demonstrate this phenomenon using continuous, non-invasive CBF monitoring via DCS.

Multiple possible physiologic reasons may explain the presence of asymmetric perfusion, though our study was not powered to elucidate specific etiologies. All four comatose subjects underwent peripheral VA ECMO, whereby blood is ejected from the ECMO circuit retrograde up the aorta via the femoral artery. ECMO blood then meets intrinsic cardiac outflow in a mixing cloud^25^. Increased cardiac contractility pushes this cloud distally, supplying the brain with native cardiac blood via the carotid arteries which diverge from the proximal aorta. On the other hand, increased ECMO flow pushes the cloud proximally toward the heart, perfusing the brain with ECMO circulation. Native and ECMO blood typically have different pH levels, oxygen saturations, and amounts of dissolved carbon dioxide. If the mixing cloud lies in between the brachiocephalic trunk and left common carotid artery, the two sides of the brain may receive blood with different levels of oxygen, CO_2_, pulsatility, and pressure^18^. A previous study looking at angiography of the head in VA ECMO patients illustrated this asymmetric blood flow via filling time of injected iodine contrast^17^.

Underlying cardiogenic shock, cardiac arrest, and ARDS may predispose the brain to hypoxic-ischemic brain injury (HIBI) before ECMO is initiated, which can prime the brain for secondary injury^26^. Cerebrovascular dysregulation is a feature of HIBI^27^ and, in the setting of asymmetric ECMO blood flow, may lead to hemispheric differences in CBF regulation. In the setting of disrupted cerebral autoregulation post-arrest, optimal MAPs for cerebral perfusion are often higher than the clinically-accepted norm of 65 mmHg^27-29^. Furthermore, interhemispheric communication may be disrupted resulting in regional independence of function and thus BFI oscillations that are independent on each side of the brain. Disruption in interhemispheric communication is seen in patients with traumatic brain and concussion^30, 31^, and thus may play a role in other global brain injuries such as HIBI. In addition to this, asymmetry in ischemic damage may result in differential dysregulation of cerebrovascular coupling, or asymmetry in in the cerebral metabolic rate of oxygen consumption (CMRO_2_) and thus oxygen extraction fraction and local CO_2_ production. There are many contributing factors: cerebrovascular reactivity to vasomotor stimuli is reduced^32^, CMRO_2_ is reduced in regions of cell death^33^ and CBF is reduced in proportion to neural activity (and thus CMRO_2_)^34^.

Interestingly, our data shows there may be an increased sensitivity to the effects of post-oxygenator pH and pCO_2_ on rBF asymmetry, with significant differences in linear model slopes between comatose and non-comatose patients. However, the associations themselves were not found to be significant. This initially confusing discrepancy is likely due to low degrees of freedom secondary to small sample sizes when assessing each individual association’s significance, vs. increased degrees of freedom when comparing the two models against one another. Arterial blood gas pH and pCO_2_ obtained from the patient’s right radial artery, representing native circulation, were not correlated with ASYM_rBF_, and importantly, nor were any differences noted between the two subgroup models. As implied above, it is possible that the left side of the brain is being exposed to the post-oxygenator blood more than the right side, and thus the effects of pCO_2_ on brain perfusion and autoregulation may be asymmetric. While this evidence is not enough to claim an differential association exists, it certainly points to one and warrants further investigation. Should this trend hold true in larger future studies, it could provide a novel piece of evidence that post-oxygenator parameters should be controlled more tightly and in a more individualized manner for patients with suspected brain injury.

Previous studies have measured cerebral autoregulation using cross-correlation analysis between continuously measured blood pressure and a neuromonitoring modality, invasive (intracranial pressure, brain tissue oximetry) or non-invasive (NIRS-based cerebral oximetry, TCD-based CBF velocity)^35-38^. We did not use this method because MAP variation was low (<5 mmHg) for long periods during monitoring, thus did not yield wide enough MAP ranges to generate full autoregulation curves each day. We did not utilize a circuit manipulation protocol to challenge MAP as has been performed in other studies^39, 40^ because of the possible danger of secondary brain injury from hypo- or hyperperfusion. Though our methodology was unable to explore the upper and lower limits of autoregulation for each patient, optimal MAPs that minimized asymmetry can be identified in Figure S3. The variability of this optimal MAP may represent the variability of individual blood pressure targets for optimal cerebral perfusion observed in ECMO patients with HIBI^41^. Past clinical trials of increased MAP goals for cardiac arrest survivors resulted in improved cerebral oxygenation, but not improved outcomes^42, 43^. However, none of these studies utilized individualized, autoregulation-based MAP targets^44, 45^. The hypothesized benefit of CBF augmentation in this population is predicated on patchy hypoperfusion due to the no-reflow pathophysiology^46^. In contrast, the physiology by which asymmetric dysregulation may occur in our cohort remains to be elucidated.

Several limitations to our study must be considered when interpreting these data. Our study captured a small population with heterogeneous pathologies ranging from cardiac arrest to acute heart or lung failure. Not all patients were treated with the same modality of ECMO, and the physiology of VA ECMO differs significantly from VV ECMO as the latter relies on the native heart for forward uniform circulation. Subjects varied by other factors such as cardiac function, level of sedation, receipt of neuromuscular blockers, and severity of illness (e.g., pre-ECMO SOFA score). Our small sample size precluded the ability to analyze these factors as covariates for association between ASYM_rBF_ and GCS-M. Future studies will focus on specific ECMO populations, such as VA ECMO recipients who have experienced cardiac arrest as they have a higher pre-test likelihood for cerebral dysregulation, to control for this heterogeneity.

One challenge we faced in designing our study was the need to dichotomize patients into ‘comatose’ vs. ‘non-comatose’ groups. Fundamentally, coma is a disordered state of consciousness in which the individual has no interaction with their environment^47^. A robust clinical definition of coma in the acute setting has not yet been developed and has often been disease-specific in prior research^47^. Most of the subjects in this study required ECMO support for cardiac pathology (e.g., cardiac arrest & cardiogenic shock), thus we drew from studies in this field to guide our definition. One of the few available interventions for the prevention of neurologic injury in CA patients is targeted temperature management (TTM). Criteria to initiate TTM require that patients be unable to follow verbal commands (GCS-M score < 6)^48^. Since all subjects in our study that had GCS-M scores of 5 eventually reached a score of 6, we have effectively adopted this dichotomization criteria. We believe this to be a good operational definition as it can be performed at the beside by a variety of trained practitioners, does not require any extra equipment, and is feasible to perform in a wide variety of clinical contexts.

To derive rBF, the median BFI value was defined as the baseline for each monitoring session as was described in the method section. However, previous DCS studies define baseline differently^11, 13, 49^. In these studies, the baseline BFI is defined as a period of the measurement where the subject is at rest. This period of rest precedes a defined perturbation, allowing comparison of the relative change of BFI between the baseline and perturbation. Since this study did not include controlled perturbations, defining such a baseline period for these subjects is difficult and arbitrary. Additionally, there is no indication that the selected baseline region corresponds to when the subject is truly at rest, thus introducing user selection bias. We did not observe statistical significance in |ASYM_rBF_ | between the two groups when using this method of normalization.

Alternatively, the mean value of the BFI time series could be used as a baseline for calculating rBF^50^ but this would be disproportionately affected if there are spontaneous changes in BFI. The median is less affected by such changes than the mean. We compared the median with the mean resting baseline values calculated in the following manner. First, each monitoring period was smoothed using a moving average filter and regions where drastic flow changes occurred were removed. This modified signal was then averaged to define a baseline to calculate rBF. The difference between the median and the mean resting baseline varied case-by-case (i.e., sometimes they matched well, and other times they differed significantly). Despite the difference, ASYM_rBF_ computed using the mean resting baseline for normalization showed statistical significance between the two groups (comatose vs non-comatose) as was shown using the median value (p < 0.006 for one-tailed and p < 0.01 for two-tailed). This affirms that using the median value as the baseline definition did not alter the major findings of this study. This might be due to our data having relatively few incidents of drastic changes in BFI as many patients were sedated or kept stable. While this mean resting baseline method seems to capture the baseline well, it does require user input when deciding on regions of drastic BFI changes. Thus, we chose to use the median as the baseline to allow for consistent selection for all the measurement periods. Future studies will continue to investigate the optimal method to estimate baseline values.

Measuring blood flow with DCS also presents its own set of limitations. As with all near-infrared spectroscopic methods, DCS is limited in its depth of penetration to cortical tissue^51^. Blood flow in subcortical regions is not well-captured. Moreover, spatial resolution was limited to the forehead because DCS is much more sensitive to interference from hair than continuous wave NIRS, which has been employed with multiple detectors throughout the head in other studies^52^. The lack of spatial resolution is less likely to be a limitation in whole-brain injury, including states of reduced consciousness or coma, as the entire hemisphere may be affected to a similar magnitude^12, 53-56^.

Another limitation of our DCS measurements was the use of constant optical coefficients of scattering and absorption. These coefficients were based on data in the literature^22-24^. Incorrect values can lead to error in the BFI calculation, particularly the scattering coefficient. It is reasonable to assume the scattering coefficient does not change during brain monitoring. However, absorption may change based on the changes in total hemoglobin concentration or blood oxygen saturation. Relative changes in blood flow are less sensitive to absorption changes, compared to absolute BFI values. Our results remain valid since we are quantifying the difference between hemispheres based on relative changes in blood flow. Nevertheless, concurrently quantifying absorption and scattering coefficients will improve the accuracy of the BFI. In the future, a frequency domain near-infrared spectroscopy system will be added to extract coefficients of absorption and scattering alongside DCS measurements.

The manner in which DCS was utilized in this study does not allow for measurements of absolute blood flow, but instead uses a relative measure of CBF. Our asymmetry measure is based on the relative blood flow normalized to the median value for each day, which allows us to better compare between days and subjects. Absolute blood flow calibration can require imaging modalities such as CT perfusion^57^ or MRI arterial spin labelling^21, 58^. We decided against using these techniques for this pilot study to minimize transport to CT scanners and avoid nephrotoxic exposure to intravenous contrast dye; furthermore the ECMO circuit is not MRI compatible. A recent study utilized time-resolved dynamic contrast enhanced NIRS (DCE-NIRS) in conjunction with a one-time bolus of indocyanine green to calibrate DCS measurements in critically ill adults^59^. The use of indocyanine green may be contraindicated in patients with renal dysfunction, which is common among patients with severe cardiac disease^60, 61^. Nevertheless, absolute CBF calibration may be important in quantifying differences in CBF as the cause of asymmetric perfusion.

## Conclusion

In this study of adult ECMO recipients, comatose subjects demonstrated increased cerebral hemispheric perfusion asymmetry as measured with DCS when compared to non-comatose patients. To our knowledge, this is the first use of DCS to measure cerebral perfusion non-invasively in this population. Future studies will include larger sample sizes and other modalities of neuromonitoring to validate hemispheric asymmetry as a biomarker of neuronal dysfunction in this population. Ultimately, it remains to be seen whether this marker is a predictor of coma recovery, informs pathophysiology, and provides a goal for targeted ECMO therapy.

## Data Availability

All data produced in the present study are available upon reasonable request to the authors.

## Acknowledgements

The authors would like to thank the staff of the Cardiac Intensive Care Unit at the University of Rochester Medical Center, without whom this study would not have been possible, as well as the University of Rochester Research Award committee for providing funding.

## Author contribution statement

**TWJ** contributed to study concept and acquisition, analysis, and interpretation of the data, and played a significant role in drafting this manuscript.

**ID** contributed to study design and acquisition, analysis, and interpretation of the data, and played a significant role in drafting this manuscript.

**KD** contributed to data acquisition and analysis.

**YX** contributed to data acquisition and analysis.

**ES** contributed to data acquisition and analysis.

**OS** contributed to study concept and design, as well as data interpretation, and revised this manuscript for intellectual content.

**MAM** contributed to study concept and design, as well as data interpretation, and revised this manuscript for intellectual content.

**RKM** contributed to study concept and design as well as data interpretation, and revised this manuscript for intellectual content.

**TTW** contributed to statistical analysis.

**IG** contributed to data acquisition and study design, and revised this manuscript for intellectual content.

**GS** contributed to study concept and design, and revised this manuscript for intellectual content.

**RC** contributed to study concept, design, data acquisition, analysis and interpretation, and revised this manuscript for intellectual content.

**IK** contributed to study design, concept, data acquisition, interpretation, and drafted and revised this manuscript.

## Supplementary information

Supplementary figures and tables can be found on the JCBFM website.

## Supplemental Figures & Tables

**Figure S1:**
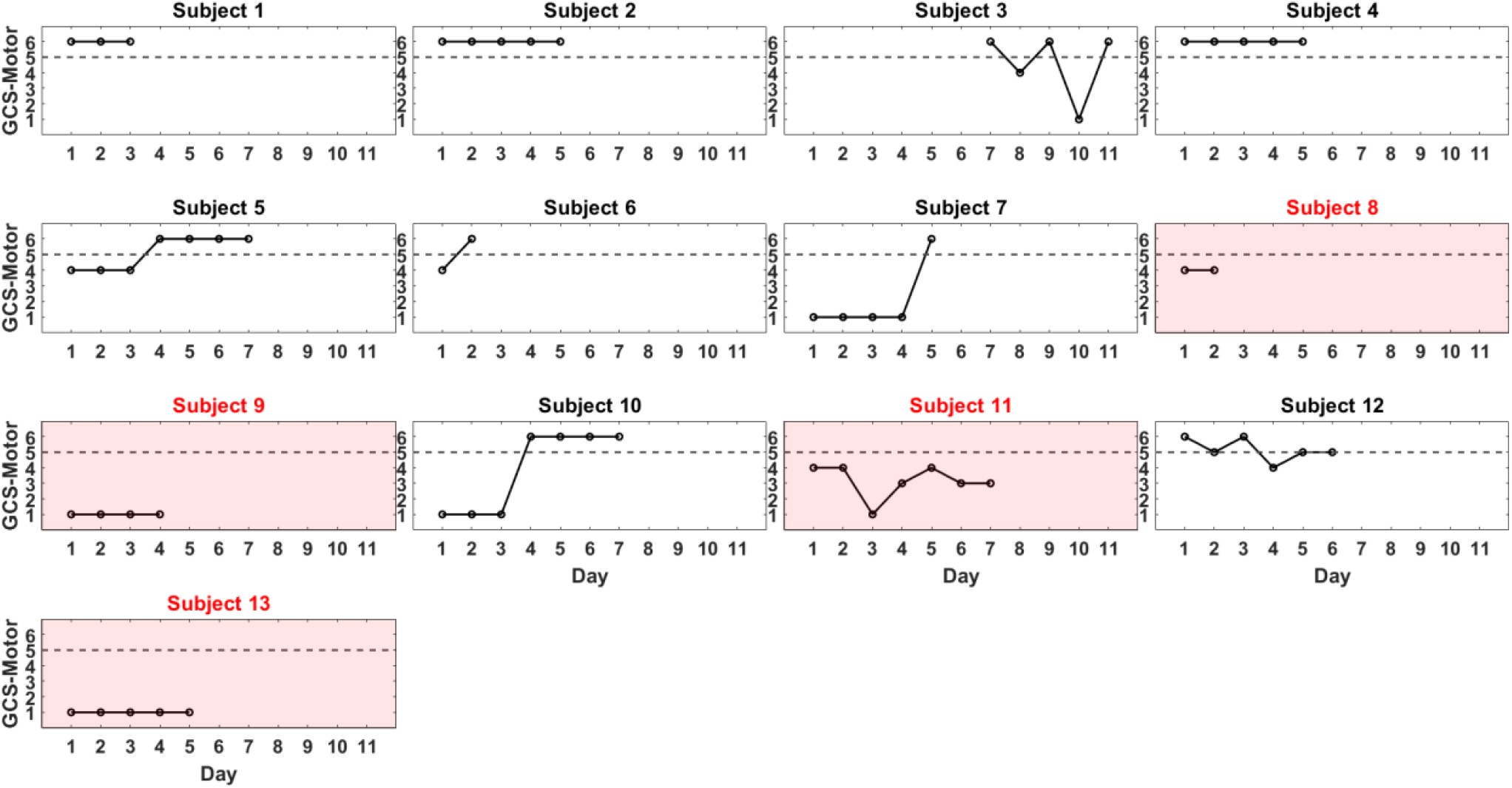
GCS-Motor scores for all the patients for each day of their monitoring. Dashed line indicates a GCS-Motor score of 5, the threshold of whether a patient is non-comatose. Subjects 7 and 10 had low GCS-Motor scores on the first days of monitoring due to paralytic medication. GCS data for the first 6 days of monitoring for subject 3 are not reported since ASYM_rBF_ was not calculated for those days. For subjects 8, 9, 11, and 13 which are highlighted in red, their GCS-Motor score never increased above 4, and thus were defined as comatose.

**Figure S2:**
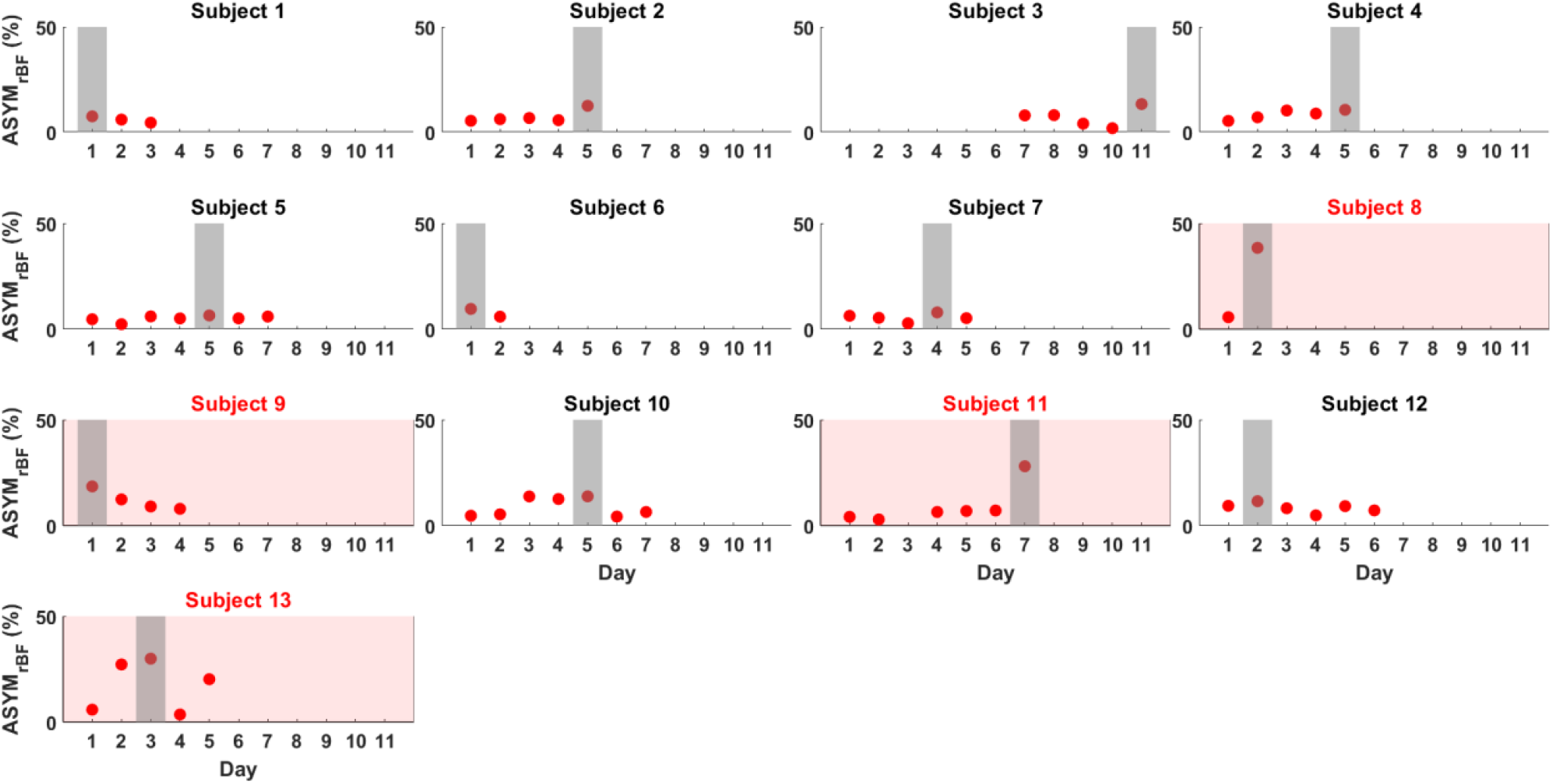
ASYM_rBF_ (red) plotted for all patients during their monitoring period. The shaded region denotes the day ASYM_rBF_ was the maximum for each subject. Data from Subject 3 days 1 through 6 and subject 11 day 3 were discarded due to low SNR. Subjects highlighted in red were subjects that were defined as comatose during ECMO treatment.

**Figure S3:**
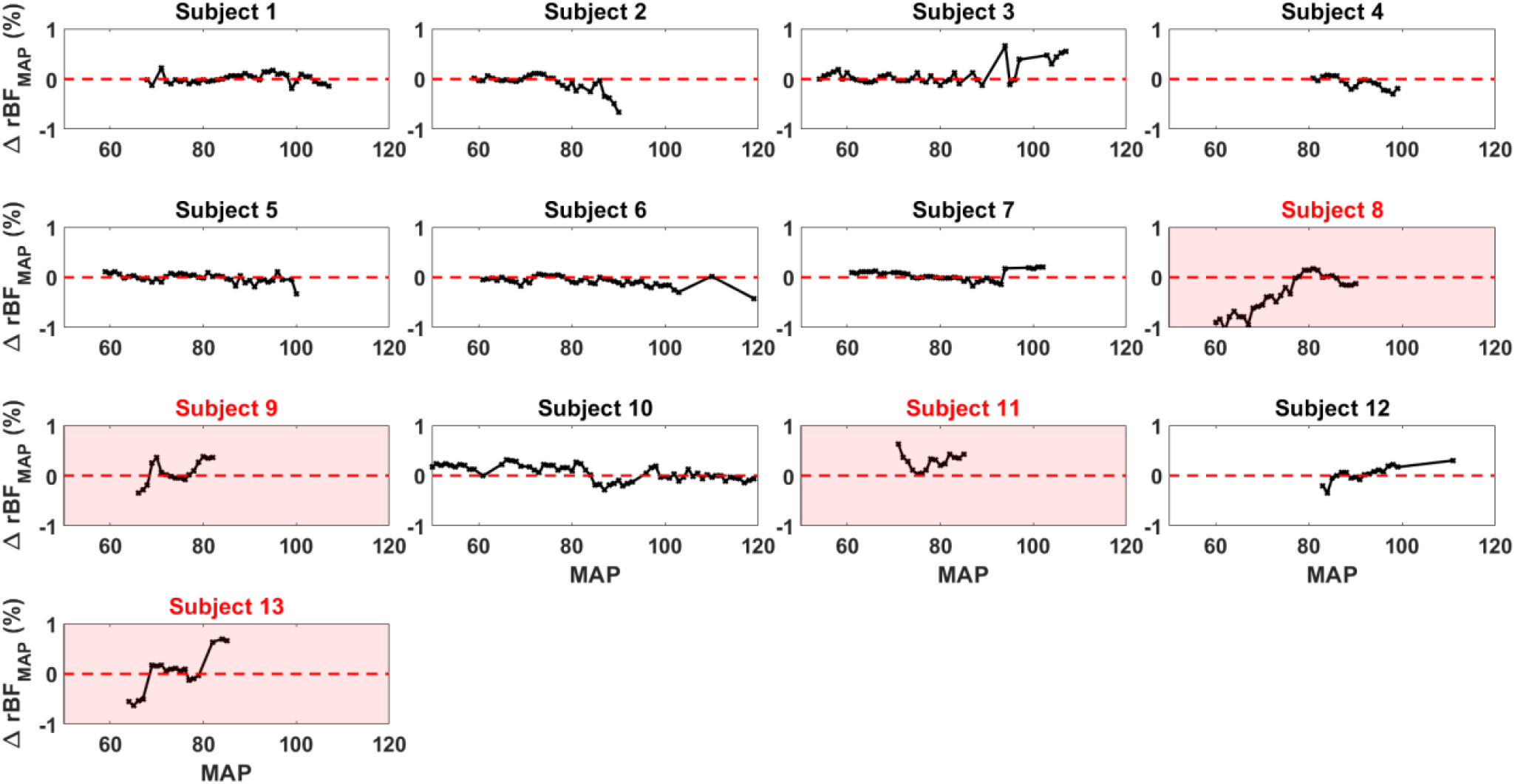
ΔrBF_MAP_ graph of all subjects. Subjects (8, 9, 11, and 13) were defined as comatose and highlighted in red. Red dotted line indicates a ΔrBF_MAP_value of zero (symmetric rBF). Negative ΔrBF_MAP_ values indicate right hemisphere rBF > left hemisphere.

**Table S1:** Average value of clinical characteristics for comatose and non-comatose patients for the day of maximum ASYM_rBF_. ABG: Arterial Blood Gas. Pre- and Post-Oxy indicate blood drawn from pre and post oxygenator of the ECMO machine.

| Clinical Variable | Non-Comatose (N=9) | Comatose (N=4) |
| --- | --- | --- |
| Monitoring Duration (hours) | 2.81 | 2.06 |
| SOFA | 12 | 10 |
| ABG Lactate | 1.5 | 1.9 |
| ABG pCO<sub>2</sub> | 37.6 | 35.5 |
| Pre-Oxy pCO<sub>2</sub> | 43.7 | 49.0 |
| Post-Oxy pCO<sub>2</sub> | 39.2 | 30.3 |

